# Associations of Disease Activity, Depression Severity, and Quality of Life in Chronic Spontaneous Urticaria

**DOI:** 10.1101/2025.09.08.25335227

**Authors:** Tuba Vural, Hacer Vural Karatoprak

## Abstract

**Introduction:** Chronic spontaneous urticaria (CSU) is often a psychophysiological comorbidity. Depression is believed to both cause and sustain CSU.

**Objectives:** We aimed to investigate the severity and presence of depression in patients with CSU and the impact of CSU on quality of life.

**Methods:** We enrolled 60 CSU and 30 healthy controls. We calculated the urticaria activity score with Urticaria Activity Score-7 (UAS-7) and divided the chronic urticaria patients into 2 groups as 30 CSU with UAS-7 below 28 (mild-moderate severity) and 30 CSU with UAS-7 28 and above (severe urticaria). We used the patient health questionnaire-9 (PHQ-9) to measure the severity of depression in all patients and the dermatology quality of life index (DQoLI) to measure the quality of life in 60 CSU patients.

**Results:** The percentages of mild depression in the mild-moderate CSU, severe CSU and healthy groups were 16.67%, 46.67%, 6.7%, respectively. The percentage of mild depression in the severe CSU group was statistically significantly higher than in the mild-moderate CSU group (p=0.0001). The number of patients with mild depression was higher in patients with mild-moderate CSU than in the healthy group, but this was not statistically significant (p=0.4238). There was a positive correlation between UAS-7 and PHQ-9, DQoLI values were r=0.545, p=0.000; r=0.941, p=0.000 respectively, and these were statistically significant. There was also a positive correlation between PHQ-9 and DQoLI, which was statistically significant (r=0.510, p=0.000).

**Conclusions:** Having severe CSU disease was closely associated with increased prevalence of depression and impaired quality of life.

## Introduction

Chronic spontaneous urticaria (CSU) is a distressing skin condition characterized by spontaneous wheals (hives), angioedema, or both, occurring without an identifiable external trigger for more than six weeks [1].

This disease affects 1% of the world’s population and has been found to greatly impair patients’ daily functions and general comfort [2].

Acute urticaria usually resolves rapidly with avoidance of the causative factors and symptomatic treatment. However, the common causes of CSU remain unidentified in approximately 70% of cases, making its management difficult and resulting in frustration for both the patient and the physician. Although the duration of CSU has been evaluated in several studies so far, it is known that it lasts for 1 year in 70% of patients and more than 5 years in 14% [3, 4]. Furthermore, CSU has been reported to significantly impact patients’ quality of life.

Persistent itching, cosmetic concerns and sleep disorders in CSU patients pose a significant burden on their physical and psychological health. The disease is increasingly recognized for its profound psychological and emotional impact beyond its physical symptoms. Numerous studies have reported an increased prevalence of anxiety and depression among patients with chronic spontaneous urticaria and that these psychological disorders parallel urticaria disease activity [5, 6]. However, it is unclear whether comorbid psychiatric conditions predispose patients to developing CSU or whether they emerge during the course of the disease. However, psychological issues may contribute to the poor QoL reported by CSU patients and may affect the outcome of urticaria treatments.

Patients’ constant itching and attacks of urticaria that cannot be stopped due to poor response to treatment increase their emotional tension.

It has also been reported that activation of mast cells in the skin during stress and their release of neuropeptides may worsen symptoms and strengthen the bidirectional link between urticaria severity and psychological health [7].

## Objectives

In this study, we aimed to investigate the relationship between urticaria disease activity (Urticaria Activity Score (UAS7)) and depression severity (Patient Health Questionnaire 9 (PHQ-9)), dermatological quality of life index (DQoLI) in patients with chronic spontaneous urticaria. A better understanding of these relationships will support more holistic and individualized patient care and enhance both the physical and psychological well-being of patients.

## Methods

### Subjects

Our study was a single center observational, cross-sectional, descriptive study. 90 people aged 18-65 years who applied to the Immunology and Allergy Diseases outpatient clinic of Isparta City Hospital were recorded. We enrolled 60 chronic spontaneous urticaria (CSU) and 30 healthy controls. We calculated the urticaria activity score with UAS-7 and divided CSU patients into 2 groups as 30 CSU patients with UAS-7 below 28 (mild-moderate CSU) and 30 CSU patients with UAS-7 28 and above (severe CSU). We used the patient health questionnaire-9 to measure the severity of depression in all patients and the dermatology life quality index to measure the quality of life in 60 CSU patients.

The inclusion criteria for the healthy group were those aged between 18 and 65 years with normal leukocyte count, C-reactive protein and erythrocyte sedimentation rate in complete blood count.

Inclusion criteria for chronic spontaneous urticaria groups were: being between 18-65 years old, having urticaria symptoms for more than 6 weeks, stopping antihistamine medications 7 days ago and not using immunosuppressive medications. Patients who only had recurrent angioedema (but did not develop wheals) (Chronic angioedema patients) and patients with urticarial vasculitis or other chronic skin diseases were excluded.

Patients were approached during standard outpatient clinic visits and were informed about the study. If they agreed to participate, they were included in the study.

This study was approved by the Clinical Research Ethics Committee of Suleyman Demirel University Faculty of Medicine (Approval number: 2025/34, Approval date: August 18, 2025). Written informed consent was obtained from each patient. The study followed the rules of the latest version of the ‘Helsinki Declaration’ and ‘Guidelines for Good Clinical Practice’.

### Demographic and clinical data

Demographic data for each patient, such as age, gender were obtained from patients through a self-reported form, and clinical data, such as disease duration, the presence of angioedema, the presence of atopy were confirmed through medical record review.

For valid assessment of CSU severity, clinicians most commonly employ the Urticaria Activity Score over 7 days (UAS7), also recognized as the gold standard in measuring disease activity.

The UAS7 was used to measure assessing two major symptoms—number of wheals, and intensity of itch—every day for seven days in succession. The symptoms are graded on 0–3 scale: 0 = None, 1 = Mild (<20 wheals/24h), 2 = Moderate (20–50 wheals/24h), 3 = Severe (>50 wheals/24h or extensive confluent areas) as wheal score. 0 = None, 1 = Mild, 2 = Moderate, 3 = Severe as itch score. The wheals and pruritus are totaled on a daily basis (up to 6 daily), resulting in a weekly score between 0 and 42. More active disease is represented by higher UAS7 score, and specific thresholds are typically used to define levels of disease control: 0–6: well controlled, 7–15: mild activity, 16–27: moderate activity, 28–42: severe activity [1].

Despite the usefulness of tools like the UAS7, they primarily capture physical disease parameters and do not account for the broader impact of CSU on patients’ lives. Therefore, additional tools are necessary to assess quality of life and emotional distress. One such tool is the Dermatology Quality of Life Index (DQoLI), developed by Finlay and Khan (1994), which evaluates the psychosocial burden of dermatological diseases. The DQoLI includes 10 items covering six domains: symptoms and feelings, daily activities, leisure, work/school, personal relationships, and treatment effects.

Dermatology Quality of Life Index (DQoLI) scores are graded as follows:

0–1: No effect at all on quality of life.

2–5: Negligible effect.

6–10: Moderate effect.

11–20: Very large effect.

21 and above: Extremely large effect

The total score can range from 0 to 30, with higher scores indicating a greater impact of dermatological issues on daily life.

### Assessment of depression level

To assess depression severity in all patients, this study used the Patient Health Questionnaire-9 (PHQ-9). The PHQ-9 is a self-administered screening tool for depressive disorder. It comprises nine items rated on a scale from 0 (“not at all”) to 3 (“nearly every day”), providing a total score from 0 to 27 [8].

PHQ-9 scores are graded as follows: 0 to 4: Minimal depression.

5 to 9: Mild depression.

10 to 14: Moderate depression.

15 to 19: Moderately severe depression.

20 to 27: Severe depression.

These scores are based on the frequency of depressive symptoms experienced in the past two weeks.

This tool is widely used due to its ease of administration and strong psychometric properties and is validated in both general medical and dermatological populations [9].

### Statistical analysis

Data were presented as mean ± standard deviation, number and percentage. In the bivariate analysis, the chi-square test were used to compare categorical variables. Mann-Whitney U test was used to compare two groups for variables that did not show normal distribution. Student T test was used to compare two groups for variables that showed normal distribution.

Fisher’s exact test was used for pairwise comparison of the percentage of depression in mild-moderate chronic spontaneous urticaria, severe chronic spontaneous urticaria and healthy groups.

Spearman Rho correlation test was used to evaluate the associations between UAS-7 and dermatology quality of life index and depression status and severity, p < 0.01 was considered statistically significant.

All analyses were performed using the Statistical Package for the Social Sciences (SPSS) version 20.

## Results

### Demographic and clinic data of mild-moderate CSU and severe CSU groups

A total of 60 CSU patients were included in this study; demographic and clinical characteristics of the participants were summarized in Table 1. In group 1 including mild-moderate (UAS-7<28) CSU patients and group 2 including severe (UAS-7>28) CSU patients, the female-to-male ratio was 1 and 7:3; the mean age was 39 ± 10.35 and 37.96 ± 13.96, respectively (p=0.114 ; p=0.715).

**Table 1.**
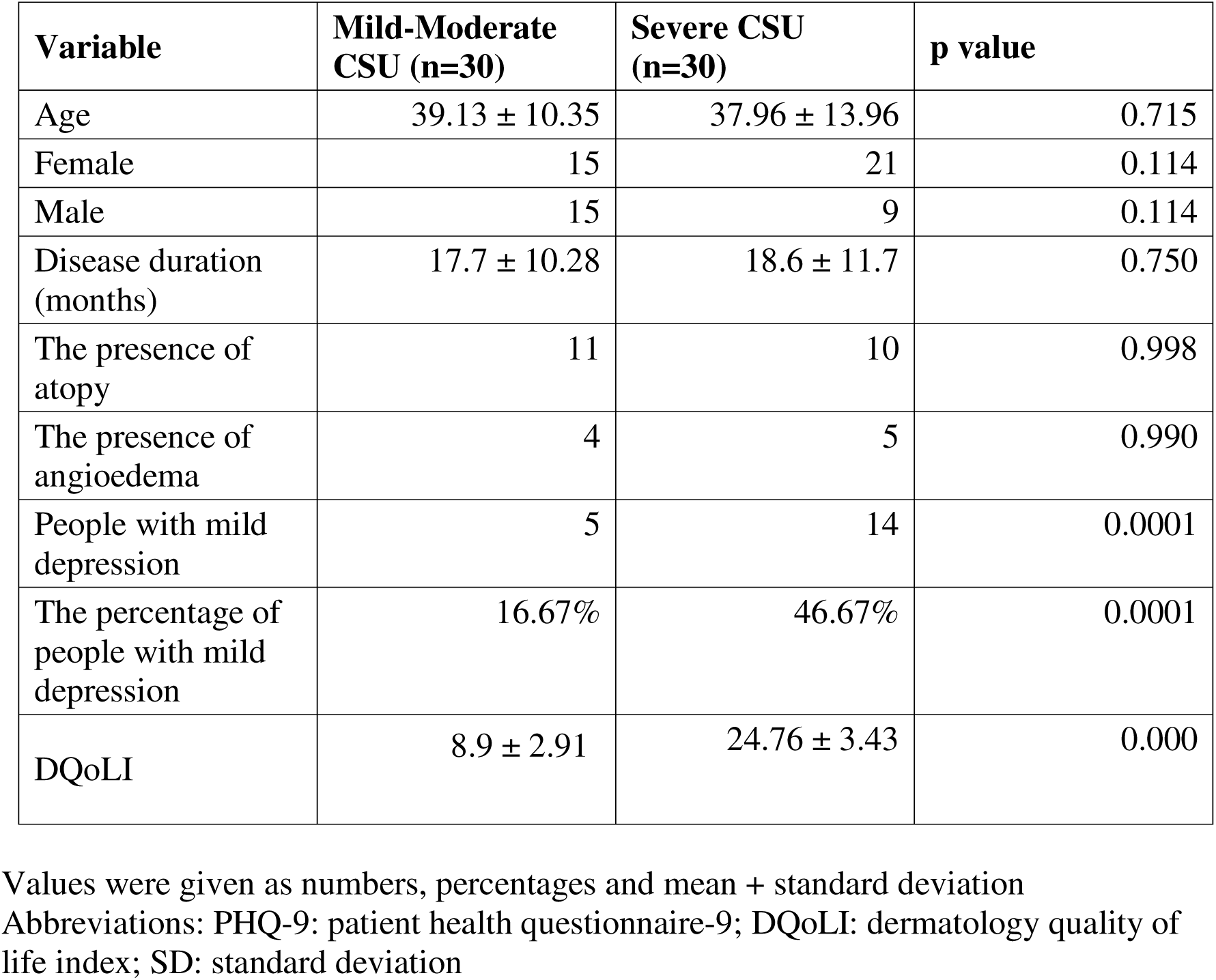
Comparison of demographic, clinical data, level of depression and dermatology quality of life index in mild-moderate CSU and severe CSU groups.

| **Variable** | **Mild-Moderate CSU (n=30)** | **Severe CSU (n=30)** | **p value** |
| --- | --- | --- | --- |
| Age | 39.13 ± 10.35 | 37.96 ± 13.96 | 0.715 |
| Female | 15 | 21 | 0.114 |
| Male | 15 | 9 | 0.114 |
| Disease duration (months) | 17.7 ± 10.28 | 18.6 ± 11.7 | 0.750 |
| The presence of atopy | 11 | 10 | 0.998 |
| The presence of angioedema | 4 | 5 | 0.990 |
| People with mild depression | 5 | 14 | 0.0001 |
| The percentage of people with mild depression | 16.67% | 46.67% | 0.0001 |
| DQoLI | 8.9 ± 2.91 | 24.76 ± 3.43 | 0.000 |

Values were given as numbers, percentages and mean + standard deviation
Abbreviations: PHQ-9: patient health questionnaire-9; DQoLI: dermatology quality of life index; SD: standard deviation

The mean duration of CSU was 17.7 ± 10.28 and 18.6 ± 11.7 months in groups 1 and 2, respectively (p=0.75). The number of people with atopy was 11 (36.66 %) and 10 (33.33 %) in groups 1 and 2, respectively (p=0.99).

The number of people with angioedema was 4 (13.33%) and 5 (16.66%) in groups 1 and 2, respectively (p=0.99). All data were statistically similar between the 2 groups.

### Level of depression and dermatology quality of life index of mild-moderate CSU and severe CSU group

Depression levels and dermatology quality of life index scores of mild-moderate CSU and severe CSU groups were summarized in Table 1.

The cut-off score for depression diagnosis was accepted as 5. The highest score received in both groups was 9. Those with PHQ-9 scores between 5 and 9 were diagnosed with mild depression. The diagnosis corresponding to the highest score they received was mild depression, and the rate of those receiving this diagnosis was reported as a percentage in both groups.

The percentage of mild depression in the mild-moderate chronic spontaneous urticaria group and the severe chronic spontaneous urticaria group were 16.67% and 46.67%, respectively. The percentage of mild depression in the severe chronic spontaneous urticaria group was statistically significantly higher than that in the mild-moderate chronic spontaneous urticaria group (p=0.0001).

Dermatology life quality index scores in the mild-moderate chronic spontaneous urticaria group and the severe chronic spontaneous urticaria group were 8.9 ± 2.91 and 24.76 ± 3.43, respectively. Dermatology life quality index score was statistically significantly higher in the severe chronic spontaneous urticaria group than that in the mild-moderate chronic spontaneous urticaria group (p=0.000).

A total of 90 people participated, 60 of whom were chronic spontaneous urticaria patients and 30 of whom were healthy individuals. There were 30 people in each group, and the age, gender, and mild depression percentages among these 3 groups were summarized in Table 2.

**Table 2.** Comparison of Age, Gender and the Percentage of People with Mild Depression in Mild-moderate CSU, Severe CSU and Healthy group.

| **Variable** | **Mild-Moderate CSU (n=30)** | **Severe CSU (n=30)** | **Healthy (n=30)** | **p value** |
| --- | --- | --- | --- | --- |
| Age | 39.13 ± 10.35 | 37.96 ± 13.96 | 38 ± 12.54 | 0.84 |
| Female | 15 | 21 | 16 | 0.244 |
| Male | 15 | 9 | 14 | 0.244 |
| People of with mild depression | 5 | 14 | 2 | 0.0007 |
| The percentage of people with mild depression | 16.67% | 46.67% | 6.7% | 0.0007 |

Values were given as numbers, percentages and mean ± standard deviation Abbreviations: SD: standard deviation n: number

The mean age of the mild-moderate CSU, severe CSU and healthy groups were 39.13 ± 10.35, 37.96 ± 13.96, 38 ± 12.54, respectively. The gender ratios of the mild-moderate CSU, severe CSU and healthy groups were 1, 7/3, 1.14, respectively.

There was no statistical difference between the three groups in terms of age and gender ratio, respectively (p=0.84; p=0.244).

The percentage of mild depression in the mild-moderate CSU, severe CSU and healthy groups were 16.67%, 46.67%, 6.7%, respectively. The percentage of mild depression in the mild-moderate CSU and severe CSU groups were statistically greater than that in the healthy group (p=0.0007).

Pairwise comparisons of the number of people with mild depression in mild-moderate chronic spontaneous urticaria and healthy groups were summarized in Table 3.

**Table 3.** Pairwise Comparison of the Number of People with Depression in Mild-moderate Chronic Spontaneous Urticaria and Healthy Group.

|  | Those with depression | Those without depression | Total |
| --- | --- | --- | --- |
| Mild-moderate CSU | 5 | 25 | 30 |
| Healthy | 2 | 28 | 30 |
| Total | 7 | 53 | 60 |

The number of patients with mild depression was higher in patients with mild-moderate chronic spontaneous urticaria than that in the healthy group, but this was not statistically significant (p=0.4238).

The correlation between UAS-7 and dermatology quality of life index and depression status and severity was summarized in Table 4.

**Table 4.** Correlation between UAS-7, PHQ-9 and DQoLI.

| Variables Compared | Correlation Coefficient (r) | p value |
| --- | --- | --- |
| PHQ-9 – DQoLI | 0.51 | 0.000 |
| PHQ-9 – UAS-7 | 0.545 | 0.000 |
| DQoLI – UAS-7 | 0.941 | 0.000 |

Abbreviations: PHQ-9: Patient Health Questionnaire-9; DQoLI: Dermatology Quality of Life Index; UAS-7: Urticaria Activity Score-7

There was a positive correlation between UAS-7 and PHQ-9 and DQoLI and the values were r=0.545, p=0.000; r=0.941, p=0.000, respectively, which were statistically significant.

There was also a positive correlation between PHQ-9 and DQoLI and it was statistically significant (r=0.510, p=0.000).

## Discussion

Psychiatric factors are thought to play a role in the development and exacerbation of CSU. In this study, we aimed to evaluate the effects of the severity of chronic spontaneous urticaria (CSU) on depression levels and dermatology quality of life.

A total of 60 CSU patients were included in this study. Demographic and clinical data such as female-to-male ratio, mean age, percentage of atopy, percentage of angioedema, and disease duration were statistically similar between the two groups in group 1, which included mild-moderate (UAS-7<28) CSU patients, and group 2, which included severe (UAS-7>28) CSU patients.

When the depression status and severity were evaluated, the mild depression rate according to the PHQ-9 scale was 6.7% in the healthy group, 16.67% in the mild-moderate CSU group and 46.67% in the severe CSU group. This difference was statistically significant (p=0.0007) and showed that the depression percentage increased as the CSU severity increased [10]. However, there was no significant difference in depression percentage between the mild-moderate CSU group and the healthy group (p=0.4238). This finding suggested that only severe CSU significantly increased the risk of depression.

In terms of quality of life, Dermatology Quality of Life Index (DQoLI) scores were significantly higher in the severe CSU group than that in the mild-moderate CSU group, respectively (24.76 ± 3.43 vs. 8.9 ± 2.91; p=0.000). The findings showed a significant deterioration in quality of life as disease severity increased [11, 12].

The fact that the groups were demographically homogeneous supported that the observed differences were due to the severity of the disease.

A very strong positive correlation was found between Urticaria Activity Score (UAS-7) and DQoLI (r=0.941, p=0.000), indicating that the deterioration in quality of life progresses almost in parallel with the increase in disease activity. This result was similar to a study conducted in 2016 [13].

In yet another study, higher DLQI scores in CSU were frequently associated with higher disease activity and psychological distress [14].

A moderate positive correlation was also observed between UAS-7 and PHQ-9 (r=0.545, p=0.000), indicating a significant relationship between disease activity and depressive symptoms. In a study conducted on CSU patients, Beck Depression Inventory (BDI) was used to assess depression severity. This study found a weak but significant positive correlation (r = 0.320, p = 0.013) between depression severity and UAS-7 [10].

In one study, the relationship between Psoriasis Area and Severity Index (PASI) score and PHQ-9 and DQoLI scores in psoriasis patients was examined. A moderate positive correlation (r = 0.42, p < 0.0001) was found between PASI score and PHQ-9 score, meaning that depression severity increased with increasing psoriasis severity [15]. Similarly, in our study, there was a significant correlation between PHQ-9 and DQoLI (r=0.510, p=0.000), indicating that depression has an impact on quality of life.

In conclusion, correlation analyses revealed that CSU severity was closely related to psychological status and quality of life.

It is understood that CSU is a complex disease that is not limited to the skin but has psychological and social aspects. Especially for severe CSU patients, a multidisciplinary approach should be adopted with psychiatric evaluation and supportive therapies when necessary.

## Data Availability

All data produced in the present study are available upon reasonable request to the authors
All data produced in the present work are contained in the manuscript
All data produced are available online at

https://www.index.doi.exact.com

https://www.index.title.exact.com

https://www.index.issn.exact.com

## Acknowledgements

We, the authors, would like to express our sincere gratitude to Prof. Dr. Mustafa Ender Terzioglu for his valuable feedback and insightful suggestions during the preparation of this article.

## Notes

### Competing Interest Statement

The authors have declared no competing interest.

### Funding Statement

This study did not receive any funding

### Author Declarations

This study was approved by the Clinical Research Ethics Committee of Suleyman Demirel University Faculty of Medicine (Approval number: 2025/34 Approval date: August 18 2025).

